# Beyond Survival: The impact of birth complications on postpartum wellbeing for mothers, babies, and households in Kenya

**DOI:** 10.1101/2025.06.09.25329313

**Authors:** Laura Subramanian, Dorothy Oluoch, Michuki Maina, Edna Mutua, Lauren Bobanski, Madison Canfora, Joyline Jepkosgei, Juliet Jepkosgei, Pauline Karingu, Consolata Chesang, Justinah Maluni, Sarah Dannin, Ki-Do Eum, Mike English, Ami Karlage, Felistas Makokha, David Kimutai, Kalama Fondo, Dickens Lubanga, Katherine E. A. Semrau, Danielle E. Tuller, Sassy Molyneux, Megan Marx Delaney

**Affiliations:** Ariadne Labs, Brigham and Women’s Hospital and the Harvard T.H. Chan School of Public Health, Boston, Massachusetts, USA; Health Services Unit, KEMRI-Wellcome Trust Research Programme, Nairobi, Kenya; Nuffield Department of Medicine, University of Oxford, Oxford, UK; Department of Health, County Government of Bungoma, Kenya; Department of Health, County Government of Nairobi, Kenya; Department of Health, County Government of Kilifi, Kenya

## Abstract

Evidence is lacking on how birth complications affect postpartum wellbeing in low-resource settings, and how to support post-complication recovery in these settings. To address this gap, we conducted a mixed methods study at three county referral hospitals in Kenya to explore the impact of birth complications on mothers’ and families’ postpartum experiences and wellbeing.

We used a convergent parallel study design, including a quantitative cross-sectional survey of 120 mothers at 5-10 weeks postpartum and qualitative semi-structured narrative interviews with 52 mothers and 19 family members at 1-2 weeks and 6-8 weeks after birth. The quantitative and qualitative strands were implemented and analyzed independently, then mixed during interpretation. We found that birth complications often come as an unexpected physical, emotional and financial shock to mothers and families, with immediate impacts at delivery and lingering effects in the postpartum period. Complications have a “multiplier effect” above and beyond the typical challenges of childbirth and postpartum recovery, with ripple effects on the broader household. Mothers’ physical and emotional health are affected by quality-of-care gaps for complications in facilities, leading to subsequent post-discharge challenges. Mothers’ emotional wellbeing is closely linked with their baby’s health, feeding and growth.

Our results offer a comprehensive view of postpartum wellbeing, adding to the evidence base on post-complication experiences. Our findings suggest key intervention points to better support mothers and newborns with birth complications. These include facility process improvements at delivery to ensure quality of care for complications (pain management, patient-centered communication about complications, newborn unit access and feeding support, family involvement in care), and post-discharge linkages with community health systems and holistic postnatal care for the mother-baby dyad to support post-complication recovery.

## Introduction

Maternal mortality has dropped by one-third in the past two decades as higher-quality childbirth care has become more accessible. (1) More mothers and newborns are surviving near-miss birth complications but are still at increased risk of morbidity and mortality in the postpartum period (first 42 days after birth). (2) For every maternal death, 20 to 30 mothers suffer from childbirth-related morbidity that can impact their wellbeing (3) and their newborns’ health. (4) Birth complications for mothers and babies are common in low-resource settings, ranging from 24% of births in Sub-Saharan Africa to 44% in South Asia. (5) Consequently, the global health community needs to understand how birth complications affect mothers and families to inform best practices to support their postpartum wellbeing. (6)

Current evidence on the impact of birth complications mostly focuses on specific maternal or newborn outcomes, rather than an interrelated range of outcomes comprising wellbeing. For example, maternal complications are associated with worse postpartum physical and social function (7–9) and postpartum depression, (10) and newborn complications like low birthweight (often driven by prematurity) cause emotional distress for mothers (11–13) and are linked to poorer infant feeding and growth. (14) Comprehensive impacts of birth complications on the mother-baby dyad and household are not well documented, especially in low-resource settings. (15,16) This makes it hard to identify best practices for post-complication care that holistically meet mothers’, babies’ and households’ needs. Global guidelines like the World Health Organization’s (WHO) postnatal care recommendations describe patient-centered care to promote the mother-baby dyad’s wellbeing, (17) but do not specify additional support needed for ongoing physical, emotional, and other challenges after surviving a birth complication. (6,17) This is an area of opportunity for research to inform best practices to support post-complication recovery in low-resource settings.

We conducted a study in Kenya in 2023-2024 to help address this gap by comprehensively exploring and documenting the impact of birth complications on postpartum experiences and wellbeing. The study was designed to measure domains in the project’s birth-to-postpartum conceptual model [Fig 1], which was developed through a literature review and expert convening. Specifically, we aimed to understand how birth complications affect mothers’, babies’, and families’ wellbeing in-facility and at selected timepoints between 2 and 10 weeks postpartum. Other aspects of the study (gender and empowerment influences in the postpartum period, healthcare worker perspectives) will be reported elsewhere.

**Fig 1:**
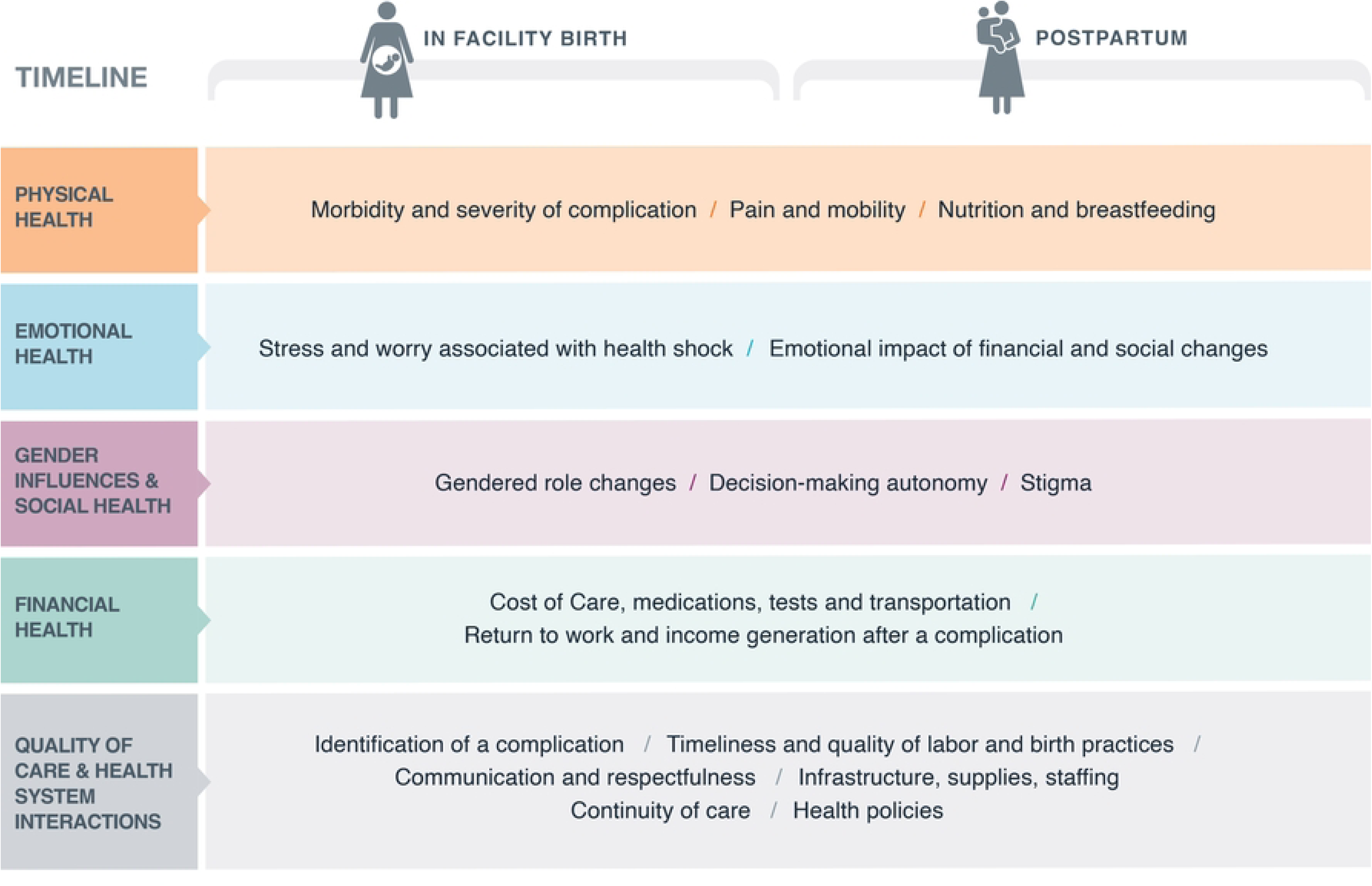
Beyond Survival Conceptual model.

## Materials and methods

### Study design

We conducted a mixed methods study using a convergent parallel design, with quantitative and qualitative strands implemented and analyzed independently, then merged at the results and interpretation levels. (18–20) We chose this design because the complementarity and triangulation of quantitative and qualitative data would allow for meta-inferences, giving us a more complete understanding of postpartum wellbeing after a complication. (18,20) The quantitative and qualitative strands had equal priority and shared grounding in the project’s conceptual model, with data collection tools designed using a matching approach to ask about concepts in similar ways across both strands. (20)

### Study setting and participants

We conducted the study at three county referral hospitals in Kenya, with catchment populations in semi-rural (coastal), rural (western), and urban informal settlement areas. The study hospitals were in the Kenya Medical Research Institute (KEMRI)-Wellcome Trust Clinical Information Network (21,22) or had other links to KEMRI-Wellcome Trust research. The site selection reflected our goal of capturing diverse intrapartum and postpartum experiences across facilities with varying geographic locations, delivery loads, and staffing, as well as cultural norms (Table 1).

**Table 1:** Study Hospital Characteristics.

|  | Site 1 (Semi-rural, coastal) | Site 2 (Rural, western) | Site 3 (Urban informal settlement) |
| --- | --- | --- | --- |
| Hospital Characteristics | Number | Number | Number |
| Annual deliveries | ~4800 | ~3000 | ~8000 |
| Annual Newborn Care Unit admissions | ~1069 | ~1188 | ~1200 |
| Neonatal cots | 21 | 18 | 13 |
| Incubators | 23 | 18 | 18 |
| Maternity beds | 51 | 50 | 65 |
| Staffing (Labor ward/postnatal) |  |  |  |
| Doctors | 1 | 9 | 24 |
| Nurses | 30 | 23 | 37 |
| Trainees | 47 | 0 | 25 |
| Neonatologists | 1 | -- | -- |
| Staffing (Newborn Care Unit) |  |  |  |
| Doctors | 0 | 6 | 9 |
| Nurses | 12 | 11 (2 per shift) | 23 |
| Trainees | 20 | 20 | 35 |
| Neonatologists | 1 | 1 | 3 |
| Nutritionists | -- | -- | 1 |

Study participants included mothers, babies, and family members. Mothers were eligible if they were aged 18+, recently gave birth at a study site, and their baby was alive at the time of enrollment and follow up. For the quantitative strand, we added eligibility criteria of singleton births (for ease of survey administration) with no urgent maternal or infant comorbidities at the time of recruitment. We prioritized enrollment of participants who experienced an obstetric complication (unplanned Cesarean section (CS) (i.e. not pre-booked), obstetric hemorrhage, eclampsia or pre-eclampsia) and/or newborn complication (preterm, low birth weight, breathing problems including birth asphyxia, sepsis). These complications are the most common and are typically unexpected and serious, making them key drivers of maternal and neonatal mortality. (23,24) We excluded planned Cesarean sections as a study complication so we could look more deeply at unique issues for unplanned health “shocks” (all subsequent mentions of “CS” refer to unplanned CS only).

Study staff determined complication status based on the reason for admission recorded in the participant’s Mother Child Health (MCH) booklet, the maternity register, and/or individual mothers’ files, with assistance from study site nurses. For comparison purposes, we also recruited some participants without study-specific complications, including uncomplicated births and those with self-reported maternal issues (e.g., anemia in pregnancy, premature rupture of membranes, breech presentation) or newborn issues (e.g., jaundice, macrosomia, birth defects). For the qualitative strand, we also included family members or close friends aged 18+ (e.g., husband, mother’s parents or in-laws, close companion) identified by participating mothers as being closely involved in their postpartum care. The recruitment period for the study began on 10-11-2023 and ended on 26-03-2024.

### Data collection methods

#### Quantitative cross-sectional survey of mothers

We developed a quantitative survey tool (S1 Fig) to measure key variables on participant demographics and pregnancy, delivery, and postpartum experiences, guided by our conceptual model. We drew on validated or field-tested measures where possible, including the Kenya Demographic and Health Survey, (25) WHO’s Maternal Women’s Voices (WOICE) Tool, (26) the 13-item Patient-Centered Maternity Care Scale (PCMC), (27) the 12-item WHO Disability Assessment Schedule (WHODAS 2.0), (28) and the Patient Health Questionnaire-2 (PHQ-2) (29) and Generalized Anxiety Disorder-2 (GAD-2). (30) We developed new questions for key concepts (e.g., mother-baby separation in facility, health self-rating) that had no existing measures but were important for understanding wellbeing. We aimed to field test the survey tool in this study, with the goal of sharing a revised tool for others to use.

We conducted two rounds of cognitive interviews with 33 mothers in January-February 2024 to test whether selected survey questions (primarily those not previously field-tested in Swahili) captured the intended constructs and could be understood and answered without difficulty. We used a reparative approach (“find and fix”) for the cognitive interviews (31) using proactive scripted probes for questions where we anticipated issues. We analyzed cognitive interview summary notes to identify patterns of interpretation, response process errors, and other problems in question administration and comprehension. The results helped us refine survey questions, interviewer instructions, and response options. For example, questions on birth complications were well understood, but spontaneous responses were more useful than probed ones, and the response options needed refinement.

We administered the survey to 120 mothers in March 2024 at the three study sites in parallel. We used a convenience sampling approach to recruit mothers (40 per site) attending study site MCH clinics at 5 to 10 weeks after birth. Clinic nurses assisted study staff to identify and preferentially recruit participants who had experienced birth complications, with efforts to include the range of maternal and newborn complications specified in the study protocol. We determined the sample size based on guidance about the necessary sample size to adequately field-test a survey, (32,33) as well as our goal of conducting descriptive analyses, with consideration of our available resources and timeframe. We administered surveys in person at the study sites using REDCap electronic data capture tools hosted at KEMRI. (34,35) 87 participants chose survey administration in Kiswahili and 33 chose English. Surveys took 30-45 minutes to administer. Respondents who screened positive on the GAD-2 (for anxiety) or PHQ-2 (for depression) were offered referrals for services.

#### Qualitative semi-structured narrative interviews with mothers and family members

We used a purposive maximum variation sampling strategy (36) to recruit 52 mothers for narrative interviews (similar numbers per site), which are designed to elicit stories of people’s experiences in their own words, allowing them to construct meaning and highlight what they consider important. (37) Mothers recruited for interviews were separate from those recruited for the survey. Mothers were recruited after birth at facilities prior to discharge, with support from nurses to identify eligible mothers and babies. We aimed to select mothers from diverse economic and cultural backgrounds, age and parity, with a range of maternal and newborn complications specified in the study protocol. Recruitment of mothers for the narrative interviews stopped once data saturation was reached (as assessed in regular sessions with the study team to review incoming data). We also conducted purposive sampling to recruit 19 family members for interviews based on nominations from participating mothers.

We interviewed the mothers and family members twice, at 1-2 weeks and 6-8 weeks after birth. We conducted two rounds of fieldwork per site between November 2023 - May 2024. The study team conducted interviews in Kiswahili in participants’ homes; interviews lasted 60-90 minutes. The semi-structured interview guides covered topics aligned with domains in our conceptual model. Interviews were audio recorded, then transcribed verbatim and translated, with verification of all transcripts to ensure accuracy and completeness of transcription. Interviewers also developed summary narrative notes following each interview.

### Data analysis

We analyzed the survey and interview data separately, then integrated the data across key themes in the conceptual model to identify convergences and divergences.

#### Quantitative analysis

We created an analytic dataset with variables for participant characteristics, complication status, pregnancy, delivery, and postpartum experiences. We conducted scoring for psychometric scales (S1 Table) in accordance with published guidance. (38)(28)(39,40) We conducted descriptive analysis to review tabulations and mean scores for variables stratified by site and complication status. All analyses were conducted in SAS 9.4 (SAS Institute, Cary NC).

#### Qualitative analysis

We imported data into NVivo 12 (Lumivero, released 2017) for coding and analysis. JoJ, CC, JM, TN and JuJ coded the data as a team, using an inductive and iterative thematic approach (41) with intercoder reliability checks. The team developed the codebook by reviewing selected transcripts to identify emerging broad themes, collectively developing an initial code list, and adding emerging themes identified in iterative discussions. The team met frequently to discuss coding and identify and resolve discrepancies. The wider study team met monthly during the study period to discuss the findings after each data collection round; their different experiences and perspectives helped in the analysis and interpretation of findings.

#### Mixed methods analysis

We integrated the survey and interview results using a merging approach, and created joint displays of data to facilitate interpretation. (19,20) We began by developing mixed methods tables structured by themes aligned with our conceptual model domains. We added survey and interview findings for each theme, stratifying by complication status. We reviewed each theme to identify convergences and divergences between the survey and interview findings and held a multi-day workshop to discuss the integrated findings. Qualitative participant demographics and complication status were quantified in order to compare sample characteristics in the two strands.

### Ethical considerations

We obtained ethical approval for the study from the KEMRI Scientific & Ethics Review Unit (SERU protocol #4712), National Commission for Science Technology & Innovation (NACOSTI/P/25/4173345), Oxford University Research Ethics Committee (OxTREC #538-23), and Harvard University Institutional Review Board (IRB23-0855). All study participants provided written informed consent and received a copy of the signed consent form. KEMRI collected limited identifying information from study participants which was kept confidential; all other study team members worked with de-identified data.

### Reporting

We are reporting on this study per the Mixed Methods Article Reporting Standards (MMARS) from the American Psychological Association, (42) and in alignment with the Standards for Reporting Qualitative Research (SRQR) (43) and the STROBE checklist for cross-sectional studies. (44)

## Results

### Participant characteristics

Of the 129 women approached for the survey, 120 (93%) consented to participate, and all participants (40 per site) completed the survey. Of the 76 women approached for interviews, 52 (68%) consented to participate (14 in Site 1, 18 in Site 2, 20 in Site 3). Of these 52 participants, 48 (92%) completed both interview rounds; 4 were lost to follow-up after the first interview. 19 family members (7 in Site 1, 6 in Site 2, 6 in Site 3) completed interviews.

Survey and interview participants had similar demographic characteristics (Table 2). Mothers were 27 years old on average; about 1 in 6 were aged 35+, and nearly half were first-time mothers. Most (84%) were married or living with a partner, and about 7 in 10 had at least secondary education. Family members were 33 years old on average and were primarily husbands (54%) or in-laws (21%); most (68%) had secondary education or higher. Participant demographics varied somewhat by study site (S2 Table), which was expected as we purposely selected sites with varying geographic locations and catchment populations.

**Table 2:** Participant characteristics.

|  | Quantitative (n=120) | Qualitative (n=52) |
| --- | --- | --- |
| **Demographic Characteristics: Mothers and Babies** | **Mean (range) or n (%)** | **Mean (range) or n (%)** |
| Baby age (weeks) | 7.5 wks (5.1 – 10.2) | R1: 2.3 wks (0.6 – 9)<br>R2: 9.9 wks (6 – 16) |
| Baby sex |  |  |
| Male | 67 (55.8%) | 29 (55.8%) |
| Female | 53 (44.2%) | 23 (44.2%) |
| Baby weight (kgs) | 2.96 (0.94 – 4.3) | 2.91 (1.20 – 4.00) |
| Maternal age (yrs) | 27.7 (18 – 43) | 27.1 (18 – 43) |
| Older mother (age 35+) | 21 (17.5%) | 8 (15.4%) |
| First time mother | 56 (46.7%) | 20 (38.5%) |
| Number of living children | 2.1 (1 – 7) | 2.2 (1 – 8) |
| Maternal education |  |  |
| Primary or less | 29 (24.2%) | 17 (32.7%) |
| Secondary | 49 (40.8%) | 22 (42.3%) |
| Higher | 42 (35.0%) | 13 (25.0%) |
| Married | 101 (84.2%) | 44 (84.6%) |
| HH head |  |  |
| Respondent | 9 (7.6%) | 5 (9.6%) |
| Spouse / partner | 94 (79.0%) | 32 (61.5%) |
| Other family member | 16 (13.5%) | 9 (17.3%) |
| Missing | 0 (0.0%) | 6 (11.5%) |
| Ethnicity |  |  |
| Luhya | 41 (34.2%) | 20 (38.5%) |
| Mijikenda/Swahili | 18 (15.0%) | 2 (3.8%) |
| Kikuyu | 14 (11.7%) | 0 (0.0%) |
| Luo | 14 (11.7%) | 5 (9.6%) |
| Other (Kalenjin, Kamba, Kisii, Meru, Somali, Taita/Taveta, Bukusu, Giriama, Other) | 33 (27.5%) | 25 (48.1%) |
| Missing | 0 (0.0%) | 4 (8.3%) |
| Worked for pay / income before birth | Worked for pay in past 12 months: 69 (57.5%) | Had income before birth: 36 (69.2%) |
| HH financial status in past 12 months |  | Not asked |
| No financial difficulties | 34 (28.3%) | -- |
| Some financial difficulties | 66 (55.0%) | -- |
| Significant financial difficulties | 20 (16.7%) | -- |
| Means of transport to facility |  | Not asked |
| Private motorized vehicle (car/ truck, motorcycle, scooter, Tuk-Tuk) | 69 (57.5%) | -- |
| Public bus | 33 (27.5%) | -- |
| Walking | 18 (15.0%) | -- |
| Time to facility |  | Not asked |
| <15 min | 30 (25.0%) | -- |
| 15-30 min | 34 (28.3%) | -- |
| 31 min – 1hr | 31 (25.8%) | -- |
| >1hr | 25 (20.8%) | -- |
|  | **Quantitative: N/A** | **Qualitative (n=19)** |

| **Demographic Characteristics: Family members** | **Mean (range) or n (%)** | **Mean (range) or n (%)** |
| --- | --- | --- |
| Age |  | 33.2 (24 - 40) |
| Relationship |  |  |
| Husband |  | 10 (52.6%) |
| In-law (mother-in-law or sister-in-law) |  | 4 (21.1%) |
| Other family member (aunt, cousin) |  | 2 (10.5%) |
| Friend or neighbor |  | 2 (10.5%) |
| Education level |  |  |
| Primary or less |  | 5 (26.3%) |
| Secondary |  | 4 (42.1%) |
| Higher |  | 5 (26.3%) |
| Missing |  | 1 (5.3%) |
| Employment status |  |  |
| Employed |  | 14 (73.7%) |
| Student |  | 2 (10.5%) |
| Unemployed |  | 2 (10.5%) |
| Missing |  | 1 (5.3%) |

As intended in our sampling approach, most study participants had birth complications (Table 3). About half (52%) of survey participants and two-thirds (67%) of interview participants had a maternal complication, mostly unplanned CS. One third (29%) of survey participants and nearly half (46%) of interview participants had a newborn(s) with a birth complication, mostly breathing problems and low birth weight. One third of survey participants (33%) and nearly half (46%) of interview participants had a baby admitted to the Newborn Care Unit (NBU), which primarily provides specialized care for sick and preterm newborns (though also provides temporary care for healthy babies while mothers are recovering from complications). 18% of survey participants and 29% of interview participants had a complication for both the mother and baby.

**Table 3:** Birth complication status of study participants.

|  | Quantitative (n=120) | Qualitative (n=52) |
| --- | --- | --- |
| **Birth Complication Status** | **Mean (range) or n (%)** | **Mean (range) or n (%)** |
| **Complication status (one or more complications listed as eligibility criterion in study protocol)** |  |  |
| None (includes "other" self-reported complications not listed in study protocol) | 45 (37.5%) | 8 (15.4%) |
| Mother only | 40 (33.3%) | 20 (38.5%) |
| Baby only | 13 (10.8%) | 9 (17.3%) |
| Mother + baby | 22 (18.3%) | 15 (28.8%) |
| **Any maternal complication (unplanned CS, obstetric hemorrhage, and/or PET)** | 62 (51.7%) | 35 (67.3%) |
| **Maternal complication type (mothers could have more than one)** |  |  |
| Unplanned Cesarean section* | 43 (35.8%) | 24 (46.2%) |
| Pre-eclampsia/eclampsia (PET) | 22 (18.3%) | 9 (17.3%) |
| Obstetric hemorrhage | 17 (14.2%) | 9 (17.3%) |
| Other self-reported complication not listed in study protocol (planned CS, sepsis, PROM, anemia, surgical complications, breech, other) | 25 (20.8%) | 1 (1.9%) - Planned CS |
| None | 33 (27.5%) | 17 (32.7%) |
| **Any baby complication (low birthweight, preterm, breathing problems, sepsis)** | 35 (29.2%) | 24 (46.2%) |
| **Baby complication type (mothers could have more than one)** |  |  |
| Low birthweight | 25 (20.8%) | 14 (26.9%) |
| Preterm | 22 (18.3%) | 9 (17.3%) |
| Breathing problems (incl. asphyxia) | 33 (27.5%) | 10 (19.2%) |
| Sepsis | 18 (15.0%) | 5 (9.6%) |
| Other self-reported complication not listed in study protocol (jaundice, macrosomia, birth defect) | 26 (21.7%) | 3 (5.8%) - Jaundice |
| None | 59 (49.2%) | 28 (53.8%) |
| **Baby admitted to NBU** | 39 (32.5%) | 24 (46.2%) |

### Impact of birth complications on in-facility experiences and wellbeing

Birth complications affected multiple aspects of mothers’ and families’ experiences in facilities, with implications for postpartum recovery and wellbeing. These impacts are described below, with additional detailed findings shown in S3 Table. Survey and interview findings on these impacts generally converged, with a few exceptions, and were similar across sites.

#### Impacts of quality-of-care gaps for birth complications

Birth complications were an unexpected shock for many interview participants, except for a small number (n=5) whose complications were identified during pregnancy. Most interview participants and nearly all (98%) survey participants had attended antenatal care (ANC).

However, the timing and number of ANC visits varied, and ANC quality gaps (health worker shortages, information gaps) and lack of money for crucial diagnostic tests like ultrasounds led to missed complication identification and preparedness.

> *“…I was told the [blood] pressure was high, that’s it, but I wasn’t told what to do to lower it, or what food to eat to lower it.”* **(Mother C2_18, maternal complication)**
>
> *“…I just thought I would give birth normally. I was very shocked to be told that I should take off my clothes and wrap myself in a cloth and go to theatre for CS…”* **(Mother C3_13, maternal complication)**

Some interview participants reported delays in care for complications during labor and delivery. These included gaps in efficiency and coordination of referrals from lower-level facilities, and long waiting times for procedures like ultrasound, blood transfusion and CS at study hospitals.

> *“It took about an hour and they were taking a patient to H5 [another facility], so we had to wait first. They took that one first to H5 and then they came and took me.”* **(Mother C1_01, maternal complication)**
>
> *“They said, ‘Tomorrow she’ll be transfused, or if we find it [blood] now, she’ll be transfused.’ Then, when you go back the next day, they say, ‘We still haven’t found it.’”* **(Family member C2_14, maternal complication)**
>
> *“I got to XXXX County Hospital at…almost 7:00 [AM]. Now I stayed for some time… I entered the theatre at about 2:00 [PM], but in between that … I was just there hurting.”* **(Mother C2_06, maternal complication)**

Several mothers reported receiving care from trainees (students); some mothers found this helpful to address staffing gaps, while others described issues with complications care from trainees (e.g., IV insertion, poor suturing) that affected their care experience.

> *“Other [trainees] used to come and work well, but there was one who came and injected me wrongly (starting an IV). He took it out again, put it in again a second time, and the water didn’t flow, now I felt pain; now he had to call another doctor and he came and took care of me…”* **(Mother C2_06, maternal complication)**

Gaps in communication regarding complications, including consent processes for treatments and medical procedures (like CS), were a key concern for mothers. Interview participants described receiving minimal information about their complications and the rationale for interventions like CS, transfusion, oxygen, IV fluids, or use of technology. They felt these communication gaps limited their decision-making autonomy for interventions and caused stress. Mothers also reported communication gaps at discharge regarding CS wound care, blood pressure (BP) management, and follow-up appointments – leaving them and their family members unsure about next steps in their recovery.

> *“There is no decision-making [for the mother]… it’s not like you’re told what to do at this hour or even the theatre [CS] consent, you’re just told, “Sign up,” just sign, you don’t even know what you’re signing up for, you see?…”* **(Mother C1_14, maternal complication)**
>
> *“I couldn’t understand what the nursery [NBU] was and what was there, but I have seen it myself now, but they don’t explain it. You see the children had a transfusion and you were not informed that the child needed blood… and if you ask what the problem is, there is a sister there who is very strict until now you are just afraid of her…”* **(Mother C2_12, newborn complication)**
>
> *“When I arrived here [home], I had those questions [how to handle the sutures and when to return for review], and I wondered why I didn’t ask about returning to have the stitches removed.”* **(Mother C1_01, maternal complication)**

Despite these challenges, as well as general quality of care issues mentioned by several mothers (disrespectful care, crowding, medication stockouts, etc.) most mothers reported their overall care experience as positive, regardless of complication status. Satisfaction with care averaged 95% for survey respondents, with a mean PCMC score of 30.6 out of 39 (higher scores indicate better care experience, with no score cutoffs or categories), with no clear pattern by complication status. Similarly, interview participants described general satisfaction and gratitude for the care they received.

> *“While I was there, the care was not bad. They just treated her well… I didn’t see anything bad. She was just treated well, and we left healthy, with the children also healthy.”* **(Family member C1_03, uncomplicated birth).**
>
> *“I can’t say the service was bad, it was just good. Because I’ve just come out fine, yes, I’ve been in pain, but that’s just a case of continuing to heal slowly.”* **(Mother C2_06, maternal complication)**

#### In-facility physical, emotional, and social support impacts of birth complications

Maternal birth complications (especially CS) had an immediate impact on mothers’ physical function and mobility, with effects sometimes lingering after discharge. Several interview participants reported significant post-CS pain, aligning with survey findings: 35% of mothers with complications had severe or extreme pain at discharge, versus 20% of mothers with uncomplicated births.

> *“The pain was unbearable [after the CS]; I couldn’t even turn over. When you wanted to get up, you’d just cry. I would just pray in my heart, asking God to help me get up so I could nurse my baby, even if just a little…”* (**Mother C3_19, maternal complication)**

Mothers with complicated births were often separated from their babies, and wished for more updates on their baby’s condition, better orientation to NBU procedures, and support with feeding their babies. Mothers described this separation as stressful and hindering breastfeeding initiation and milk production -- sometimes leading to ongoing breastfeeding challenges. In contrast, most mothers with uncomplicated deliveries started breastfeeding soon after delivery and remained with their babies throughout their facility stay. Survey findings were similar for mother-baby separation in facilities: 95% of mothers with uncomplicated deliveries got to see and hold their baby continuously or most of the time in the facility, vs. 36% of those with both mother and baby complications.

> *““He was taken to the nursery, and I was attended to and was taken to the ward, I was told, ‘Your child is in the nursery …Once you have strength, you can walk, you will have a chance to go to the nursery and see your child’… It was just stressful because your head is full, you can’t walk there, and you’re thinking about your child, you haven’t seen him.”* (**Mother C3_01, maternal and newborn complication)**
>
> *“Without seeing the child, I didn’t know where the child was. I thought they were lying to me; maybe he got there and died. When the child was taken out, he didn’t cry…I thought that they wanted me to stay for two or three days before they break the sad news of his demise.*” [Note: the baby survived] **(Mother C3_14, maternal complication)**
>
> *“Now when it comes to expressing milk, I’ve never seen it before. That expressing, I try to squeeze the milk but it does not come out and I am told to breastfeed, I struggled.”* (**Mother C3_09, maternal and newborn complication)**

Mothers recovering from complications were further stressed by lack of help from family members with baby care, driven by strict visitation policies in postnatal wards and the NBU.

> *“[After delivery]… I was just trying to manage by myself I wished my mother could have stayed with me, but whenever she came, she was asked to leave.”* **(C2_WN_15, maternal complication)**

#### In-facility financial impacts of birth complications

The financial shock of birth complications resonated throughout mothers’ facility stays, setting the stage for financial stress after discharge. Per interview participants, government health insurance schemes covered most childbirth care costs but not additional diagnostics (like ultrasounds), treatments, or extended stay expenses for birth complications. Survey participants with complications stayed longer at facilities (up to 8.7 days for mothers [range 0 – 60] and 14.2 days for babies [range 0 – 60] vs. 1.8 days [range 0 – 10] for uncomplicated births), and paid for supplies or medications more often (50% among mother + baby complication vs. 11% of uncomplicated births). Interview participants reported that out-of-pocket expenditures for complications negatively impacted families’ financial situations, putting them at a disadvantage for adhering to postpartum medications and treatments.

> *“They tell me that I should continue with those drugs and not try to stop taking them. I told them, ‘Here you are giving me free and if I go home, who will be giving it to me?’ … They prescribed five medicines. That was given to us for free, and the others are for purchase, I still haven’t bought the three medicines and I don’t even know if I will buy them: there’s no money.”* **(Mother C3_20, maternal and newborn complication)**

### Impact of birth complications on postpartum experience after discharge

Birth complications had lasting impacts on mothers’ physical and emotional wellbeing in the first few weeks postpartum, with broader effects on the mother-baby dyad and broader household. These impacts are described below, with additional detailed findings shown in S4 Table. Survey and interview findings on these impacts generally converged, with a few exceptions, and were similar across sites.

#### Impact of birth complications on mothers’ physical wellbeing after discharge

Mothers with complications (especially CS) faced ongoing physical challenges after discharge. Survey participants with maternal complications had worse physical function at 5 – 10 weeks postpartum (mean WHODAS scores up to 20.9 out of 60 [range 12 – 37], with lower scores indicating better function) than those without complications (mean WHODAS score 14.8 [range 12 – 26]), and CS was common among mothers with the poorest physical function. About one quarter (27%) of survey participants with maternal complications sought postpartum treatment for illness or specialty care (i.e., not a regular check-up) in the past 30 days, vs. 9% of uncomplicated births. Three quarters (73%) of survey participants with mother + baby complication rated their health in the past 30 days as ‘good or very good’, compared to 95% of uncomplicated births. Similarly, interview participants with complications reported ongoing challenges with physical function that impacted their wellbeing and productivity. These included post-CS challenges with wound healing and pain, and other physical symptoms like feeling unwell, dizziness, numbness, back pain, and unstable blood pressure.

> *“I’m having problems, I am praying tomorrow that if I’m able to go to the higher-level hospital, I’ll tell the doctor because my waist is paining … If I do a lot of washing or try to carry a jerrican of water for a distance, I feel pain here. Then the numbness also hasn’t stopped. I can still feel it at the CS stitches, even if you press it like this, you will feel it.”* **(Mother C1_04, maternal complication)**
>
> *“The challenges are related to pain, you can find someone to help with the chores, but sometimes not, I have to strive by myself. For example, doing laundry, if there is no one to help, you have to strive yourself.* **(Mother C1_07, maternal and newborn complication)**

Interview participants reported that postnatal care (PNC) services mainly focused on the baby, e.g., weight check and vaccines. This meant missed opportunities for mothers to talk about their own post-complication recovery, including the issues described above with wound healing, pain, and other physical symptoms. Mothers also reported that financial barriers to accessing PNC led them to seek care for themselves at dispensaries or not at all, although this was not specific to those with complications. Similarly, survey participants reported access to care barriers like getting money for treatment (32% “big problem”) or not wanting to go alone to the facility (24% “big problem”), but these barriers did not vary by complication status.

#### Impact of birth complications on mothers’ emotional wellbeing and social support after discharge

Mothers struggled with emotional wellbeing after discharge, but this was mostly characterized by general stress and worry rather than clinical postpartum depression (PPD) or postpartum anxiety (PPA) and was not clearly linked to complication status. PPD prevalence (PHQ-2 score of ≥3.0 out of 6) was low (5%) among survey participants, and most interview participants did not mention depression symptoms. Mean PHQ-2 scores were higher (worse) among survey participants with newborn complications (1.23) and maternal complications (0.63) vs. uncomplicated births (0.38), but there was no clear pattern in PPD prevalence by complication status. PPA prevalence (GAD-2 score of ≥3.0 out of 6) was relatively low (11%) among survey participants, with higher (worse) mean GAD-2 scores among those with newborn complications (1.31) and maternal complications (0.80) vs. uncomplicated births (0.62), but no pattern in PPA prevalence by complication status. Few interview participants reported clinical anxiety symptoms, but many described feeling generally stressed and worried about breastfeeding challenges, concerns about their baby’s weight gain and illness, and challenges with support during their recovery – although with no clear pattern by complication status.

> *“You are stressed up, you can’t be at peace even eating, with the baby being sick, you are just stressed up and wondering, what is the issue? Will he get better? So, you can’t be at peace.”* **(Mother C1_03, uncomplicated birth)**

Mothers’ emotional wellbeing was affected by community beliefs about birth complication causes and recovery. Interview participants noted that mothers are blamed for complications, which are believed to be associated with bad omens, witchcraft, infidelity, and extensive contraceptive use. Community beliefs also affected how much social support mothers received after discharge. Some interview participants reported that mothers with complicated births typically receive support for longer. Others described unrealistic expectations for CS recovery and return to productive duties, leading to returning to household chores too soon, struggling with those chores and feeling like a burden.

> *“Their journeys, most of them, they just said they were okay. But now, I was the only one who underwent a C-section among all the grandchildren, so everyone was worried… They were telling me I was cowardly.”* **(Mother C2_15, maternal complication)**
>
> *“Where I come from, XXX County, after giving birth, you are taken care of in everything; your only task is to bathe and rest, at least for one month. But here, on the first or second day, you’re expected to cook, and wash dishes…So, that’s something I’m trying to adopt here … even though I still feel some pain, you are expected to do it.”* **(Mother C3_16, maternal complication)**
>
> *It’s just that, that is, I’m not used to sitting, now I see it as a burden to tell someone to do this for me, to do this.”* **(Mother C1_01, maternal complication)**

#### Impact of birth complications on babies’ health and feeding after discharge

Survey and interview findings diverged on babies’ health after discharge. Nearly half (46%) of babies with complications had post-discharge admissions for illness, vs. 22% of uncomplicated births. Survey participants with newborn complications also rated their baby’s health lower in the past 30 days (77% ‘good or very good’) than others (93% - 96%). In contrast, several interview participants described baby illnesses like jaundice, respiratory infections, and GI issues that required medical treatment after discharge, regardless of the baby’s complication status.

> *“He was having difficulties breathing and wasn’t breastfeeding well. So, they prescribed some medicine, and I bought it for him.”* **(Mother C3_11, uncomplicated birth)**
>
> *“She has had these infections on the neck and ears. She had wounds all over the neck and ears with some discharge.”* **(Mother C3_07, maternal and newborn complication)**

Breastfeeding was nearly universal in the first few weeks postpartum, regardless of complication status; 85% of survey participants and many interview participants were exclusively breastfeeding their babies. Interview participants noted that newborns with complications initially needed tube- or cup-feeding in the NBU but were able to transition to breastfeeding. Some mothers introduced supplemental foods like cow’s milk, processed milk, porridge, orange juice, or water. Reasons for supplemental feeding included perceived inadequacy of breast milk (attributed in some cases to stress), insufficient baby weight gain, other breastfeeding challenges, advice from health workers or family members, or to treat constipation or colic.

There was no clear pattern of breastfeeding challenges or supplemental feeding by complication status.

> *“…I didn’t have enough milk. So, I asked my mother, and she said I would have to add a little cow’s milk, but that milk must be fresh, and that’s what I did, I give it to [him] sometimes. But I haven’t given him water.”* **(Mother C1_11, uncomplicated birth)**
>
> *“She usually drinks her mother’s milk, I sometimes give her fruits, I give her oranges and water too. She has not started drinking milk.”* **(Mother C1_12, maternal complication)**

#### Financial impacts of birth complications after discharge

Households continued to face financial impacts of birth complications after returning home. Up to 46% of survey participants with complications (vs. 31% of uncomplicated births) said their family’s financial conditions worsened after delivery. Up to 30% (vs. 16% of uncomplicated births) had to borrow money or sell property / possessions to pay for treatment related to the birth, and up to 30% (vs. 11% of uncomplicated births) did not have enough food or money to buy food in the past 7 days. Most interview participants reported some financial postpartum stress due to reduced household income and baby care expenses, with birth complications worsening this stress. Mothers reported that birth complications often led to ongoing costs for follow-up care, and it was financially challenging to comply with health worker recommendations to continue complication-related medications (e.g., baby supplements and BP meds).

> *“Even if you show them the [government insurance] card, it doesn’t help. It helps when you have given birth, and the child is not sick, that’s what they tell you. But when the child is sick, it’s a different story. There, you pay if the children are sick; the card doesn’t cover them.”* (**Mother C1_08, maternal complication)**
>
> *“Also, those medications, those medicines come at a very high cost. You might run out of the medicine and … funds have [been] delayed [not get the money to replenish]. So, you see, you might find yourself not being able to give him the supplements regularly.”* **(Mother C2_12, newborn complication)**

Interview participants noted that these additional financial constraints affected their ability to access essential care and food for themselves and their families, as well as their emotional wellbeing and family dynamics.

> *“We spent a lot of money during my operation, so I don’t want to burden my husband with so much.”* **(Mother C3_19, maternal complication)**
>
> *“So, it’s hard when you have to ask someone for help… it can feel like you clash with him. Plus, I’m used to earning my own money and doing small jobs. Now, I feel like I’m a burden to my husband. Sometimes I feel like I’m putting too much pressure on him, and it seems like he’s getting tired like he’s frustrated.”* **(Mother C3_12, maternal complication)**

## Discussion

This study leverages integrated survey and interview data to offer a comprehensive view of postpartum wellbeing after birth complications. Our findings show that complications have a “multiplier effect” on the typical challenges of childbirth and postpartum recovery, bringing immediate physical, emotional, and financial impacts that linger in the postpartum period.

Quality of care, finances, and social support all play a role in complication experiences and recovery.

We found that birth complications are often an unexpected shock for mothers and families, and while they are satisfied and grateful for the care they receive (consistent with prior studies on maternity care satisfaction (27,45,46) quality of care gaps for birth complications can negatively impact mothers’ wellbeing and exacerbate recovery challenges after discharge. Gaps in post-complication pain management and care instructions at discharge can prolong the physical effects of maternal complications, leading to lingering challenges with physical health and function (WHODAS 2.0 scores) observed in our study and other studies. (7–9) Gaps in communication and autonomy (the lowest-scoring domains of patient-centered maternity care in our study and in low-resource settings more broadly (47)) are amplified for mothers with complicated births. Mothers experience added stress due to inadequate communication about complication causes and treatments for themselves and their babies and perceive a lack of autonomy in the CS consent process, which is known to be challenging (48). Delays in care and reliance on trainees (common in understaffed wards (49)), driven by resource shortages, (50) also contribute to post-complication challenges. The baby-centered nature of PNC visits means missed opportunities to support mothers in their post-complication recovery.

Our findings show that mothers’ emotional wellbeing is closely linked to their baby’s wellbeing, with complications exacerbating stress. In facilities, mothers feel worried and stressed at being separated from their newborns without adequate communication about their baby’s condition or feeding support, consistent with prior studies. (51,52)(11–13,53) After discharge, newborns with complications have a higher prevalence of hospital admissions, with corresponding stress for mothers. Mothers struggle with breastfeeding challenges and perceived milk insufficiency, including for preterm and low birth weight babies as seen in prior studies. (14) In general, mothers feel worried and stressed about their baby’s feeding, health, and growth regardless of whether they had a complicated birth. This stress is associated with poorer breastfeeding and infant growth outcomes, (54) and is not captured by the GAD-2/PHQ-2 clinical screeners used in our study -- which showed low PPD and PPA compared to documented global levels (11-19% PPD in LMIC settings, (16,55) 66% PPD / 30% PPA among mothers with life-threatening birth complications, (56) 42% to 48% PPD among mothers with newborn complications (57)). This important measurement gap could be addressed by adapting postpartum worry/stress scales from high-resource settings, i.e., the Postpartum Worry Scale-Revised, (58) Postpartum Specific Anxiety Scale (PSAS), (59) and PSAS-RSF-C for global crisis, (60) or potentially the Parental Stress Scale (61) or Kessler psychological distress scale. (62)

Our findings also shed light on how the financial and social aspects of birth complications are intertwined with postpartum wellbeing, adding nuance to prior findings that unexpected complications can strain social and financial support systems. (63–65) Treatment and follow-up costs for birth complications exacerbate the financial stress of childbirth for mothers and families, starting in-facility and reverberating throughout the postpartum period. This financial stress makes it difficult to adhere to post-complication visits and medications, affecting mothers’ and babies’ wellbeing. At delivery, facility policies limiting family visitation during hospitalization are particularly challenging for mothers with complications, who have added needs for in-facility social support. Social support remains an important contributor to wellbeing after returning home, when mothers with complications need extra support with household work and chores (some receive it, but not always for as long as they want), and some face stigma regarding complications.

### Implications and recommendations

Our findings suggest key intervention areas to improve mothers’, babies’ and families’ wellbeing after a birth complication. These interventions align with our conceptual model and the global community’s focus on respectful, family-centered maternity care offering continuity of care after discharge. In the immediate aftermath of a complication, mothers and babies could be better supported by modifying intrapartum care processes to promote successful recovery and wellbeing. These process improvements are consistent with the WHO recommendations on intrapartum care for a positive childbirth experience, (66) and could potentially be incorporated into existing improvement efforts and care processes (which is crucial, given the formidable resource constraints faced by facilities and health workers).

- **Enhance pain management for CS**: Ensure consistent provision of pain medication and pain management guidance at discharge (particularly for CS) to enhance maternal wellbeing and promote faster recovery.
- **Strengthen trainee supervision**: To help ensure quality and safety of care, adjust trainee supervision processes to prioritize situations where trainees are providing care for complications.
- **Promote patient-centered communication**: Train health workers and improve facility processes to promote communication about complications, including the rationale for medical interventions, informed consent for CS and other medical procedures, condition updates (especially for babies in the NBU), and detailed discharge instructions.
- **Improve maternal access and orientation to NBU**: Where feasible, consider more flexible NBU access policies while maintaining infection prevention and control measures, and establish more mother-friendly spaces within or adjacent to the NBU— such as private feeding or milk expression rooms and dedicated areas for kangaroo mother care (KMC). Orient mothers to ensure they understand NBU procedures and processes.
- **Provide more intensive support for feeding sick newborns**: Support mothers of babies in the NBU on how to express milk, navigate cup feeding, and transition to breastfeeding.
- **Promote social support and family involvement in care:** Consider facility visitation exceptions to engage family members not only as birth companions, but also as care partners in providing extra support for mothers and babies after a complicated birth. Strengthen family orientation to the NBU, and sensitize family members on expectations for mothers’ recovery, including the type and duration of support they will need.
- **Enhance upstream preparedness**: Improve complication readiness and detect complications early (where possible) in ANC, which can reduce maternal and neonatal mortality (67) and lessen the emotional impact of health shocks.

Once mothers and babies return home, their postpartum wellbeing could be addressed through care improvements building on the WHO recommendations for postnatal care: (17)

- **Leverage existing community health strategies**: engage community health workers (a valuable PNC resource in Kenya (68)) to manage expectations around mothers’ recovery and support, encourage mothers to prioritize their own recovery equally to their baby’s health, and identify post-complication issues and referring mothers and babies for care. Mothers and families would also benefit from enhanced linkages to social support systems, facilitated through existing government structures and supplemented by partnerships with organizations that provide targeted support for vulnerable families and children.
- **Optimize PNC at facilities** by strengthening support for the mother-baby dyad as a connected unit:

◦ <u>Align well-baby visits with mothers’ physical and emotional health checks</u> to better meet mothers’ post-complication needs, and implement feedback loops with dispensaries where mothers often seek care.
◦ <u>Promote patient-centered communication</u>: Encourage/train health workers to ask about key areas of concern for mothers, including breastfeeding and milk production (drivers of supplemental feeding), and empathize with mothers’ worry and stress.
◦ <u>Promote social support and family involvement in care</u>: Continue engaging family members about ongoing support for mothers during post-complication recovery (e.g., not starting heavy chores too soon after CS).

### Limitations

Our study sample has some limitations. Since we used a facility-based convenience sampling approach targeting mothers who had complicated births, we do not have a representative sample of postpartum women in these areas of Kenya. Survey and interview participants had similar demographic characteristics, but their recruitment points differed so they may be different in other unmeasured ways. Our study excluded mothers under age 18, but they might have unique experiences as a vulnerable group that should be considered in the future.

Our data collection tools also have limitations. The survey and interview tools were designed to ask about concepts in similar ways, but some topics like postpartum emotional wellbeing were measured in slightly different ways and depth. For the survey, some concepts did not have good existing measures so we had to create our own, which were tested in the cognitive interviews.

Using the GAD-2 and PHQ-2 in the survey rather than the more common Edinburgh Postnatal Depression Scale allowed for parsimony and avoided cultural difficulties around asking about suicidal ideation, but limited our ability to identify sub-clinical worry and stress. We chose not to ask about some sensitive concepts in this study that may be relevant for complications experiences. For example, we did not ask about violence against women, given the potential harm of being unable to meet the needs of women who reported it. We did not ask about external factors like climate-related events (e.g., floods) or political events (e.g., doctor’s strike), but these affected women’s access to care and would be worth investigating in further studies.

Our primary goal for the survey was to field test the survey tool and collect sufficient data for descriptive analyses. Consequently, the survey had a relatively small sample purposefully targeting complicated births and was not powered to make statistical inferences, so there are limits to what we can infer from the data. For example, we had limited ability to explore previously demonstrated associations between birth complications and PPD (10) and between reduced maternal function and failure to breastfeed. (8) Since the survey tool has now been field-tested, future studies could be powered to examine these associations. The interviews focused on understanding the nuances of experiences with birth complications, so were not intended to be representative of experiences in Kenya more broadly, although we spoke with participants from a variety of backgrounds. Qualitative interviews can be subjective; researchers’ interpretations could have introduced some bias, although this was mitigated though regular coding discussions and triangulating data from mothers and family members.

## Conclusions

This study provides valuable evidence on the “multiplier” effect of birth complications on several interrelated dimensions of postpartum wellbeing for mothers, babies, and families. Our results go beyond physical and emotional health to include other important aspects of postpartum wellbeing like social support and household finances. Our findings suggest key intervention points at delivery and after discharge where mothers and newborns with birth complications can be better supported, improving their experience and wellbeing. These include improving facility processes at delivery to ensure quality of care for complications (pain management, trainee supervision, patient-centered communication, NBU access, family support); linkages with community health systems after discharge to promote post-complication recovery; and enhancing postnatal care to more holistically meet the needs of both mothers and babies.

## Data Availability

The de-identified quantitative survey dataset is available in the Harvard Dataverse Repository, which is a free data repository open to all researchers from any discipline, both inside and outside of the Harvard community. The survey dataset includes all original variables as well as derived variables created for the analysis, and can be accessed along with the data dictionary at https://dataverse.harvard.edu/previewurl.xhtml?token=56e03d16-9edc-49d4-a614-c6560c6848b0. The qualitative datasets generated and analyzed during this study are not publicly available due to the nature of this qualitative research and the potential to identify respondents, but are available from the corresponding author upon reasonable request through the KWTRP's data governance committee.

## Acknowledgements

We thank the respective County health directorate and management teams and hospital management teams for their support. We thank the clinical teams and nurses in the study sites

for their support, participation and feedback. We are grateful to the mothers and family members that participated in the study. We thank the WHO for allowing us to use parts of the Maternal WOICE tool, and Patience Afulani for providing guidance on using the PCMC. We thank Rediet Bayou for helping to conduct the literature review for this manuscript, Maria Kartika for assisting with analysis of free text data in the quantitative survey, and Onesmus Onyango for assisting with data collection. This work is published with the permission of the director of KEMRI.

## Supporting information

S1 Fig. **Beyond Survival quantitative survey tool.**

S1 Table. **Scoring methods for psychometric scales in Beyond Survival quantitative survey.**

S2 Table. **Participant characteristics, by site.**

S3 Table. **Detailed mixed methods results: impacts of birth complications on in-facility delivery experience and wellbeing.**

S4 Table. **Detailed mixed methods results: impacts of birth complications on postpartum wellbeing after discharge.**

